# Interrater agreement of seizure onset and spread in scalp EEG

**DOI:** 10.64898/2026.11.20.26366721

**Authors:** Juri Kim, William K.S. Ojemann, Carlos A. Aguila, Odile Feys, Daniel J. Zhou, Madeline Stecy, Christina M. Boada, Julian Gal, Russell T. Shinohara, Kathryn A. Davis, Erin C. Conrad

**Author notes:** Corresponding author: Juri Kim, Department of Bioengineering, School of Engineering & Applied Sciences, University of Pennsylvania, Philadelphia, PA 19104, USA.

## Abstract

**Objective:** Faster seizure spread on intracranial EEG has been linked to worse surgical outcome. If contralateral spread on scalp EEG carried similar prognostic information, it could identify poor resection candidates before intracranial monitoring and direct them toward neuromodulation sooner. Testing this first requires that experts reliably annotate spread on scalp EEG. The interrater agreement for seizure spread detection on scalp EEG is unknown; prior scalp EEG reliability studies have focused mainly on seizure detection. We quantified expert agreement on the laterality, channels, and timing of seizure onset and contralateral spread.

**Methods:** We retrospectively analyzed 79 seizures from 48 patients undergoing presurgical evaluation at a single center. Three board-certified epileptologists independently marked the time and channels of onset and contralateral spread for each seizure. We measured agreement on onset laterality and spread presence with Gwet κ, channels with Cohen κ, and timing with the median absolute deviation (MAD). Inter-rater and majority-vote consensus agreement were estimated with patient-clustered CIs.

**Results:** Onset laterality showed substantial inter-rater agreement (Gwet κ = 0.73, 95% CI 0.62–0.83); most disagreement involved bilateral onsets. Contralateral spread agreement was comparable (0.63, 0.48–0.76). Channel-level agreement was higher for onset (inter-rater Cohen κ = 0.57) than for spread (0.38). Onset timing was highly consistent (median MAD 0.41 s); spread timing agreement was also high (1.99 s), but less so than onset timing (sign rank p < 0.001). Agreement did not differ across annotators or between temporal and non-temporal epilepsy.

**Significance:** Epileptologists generally agree on onset laterality and whether and when seizures spread contralaterally, although spread channels are identified less reliably. These findings support the feasibility of measuring scalp EEG spread as a candidate noninvasive marker of seizure network extent. The public annotated dataset provides a multi-expert reference standard for developing automated spread-detection algorithms.

**Key Points:**

- Epileptologists showed substantial inter-rater agreement on seizure onset laterality on scalp EEG.
- Agreement on whether seizures spread to the contralateral hemisphere was high, but agreement on which channels were involved in spread was lower.
- Raters annotated onset and spread timing consistently (median deviation 0.41 s and 1.99 s, respectively), within windows used to define early spread.
- We provide a public scalp EEG dataset with multi-expert onset and spread annotations to support automated spread-detection algorithms.

## INTRODUCTION

The success of epilepsy surgery depends on precisely localizing the seizure onset zone (SOZ) and subsequent seizure spread.^1,2^ A central challenge is reliably distinguishing a focal, resectable epileptogenic zone from a diffuse or bilateral network in which ictal activity is not confined to a single focus; unrecognized diffuse or bilateral networks are a major contributor to surgical failure.^3^

Intracranial EEG (iEEG) studies have shown that rapid contralateral seizure spread predicts poor surgical outcome, and suggests a diffuse or bilateral epileptic network.^4–6^ However, iEEG is invasive, carries procedural risk, and has limited spatial sampling.^7^ A non-invasive counterpart to this biomarker—one detectable on scalp EEG before iEEG is undertaken—would be clinically valuable. Such a marker could, in principle, identify patients whose diffuse or bilateral networks make them poor resection candidates, allowing them to proceed directly to neuromodulation device therapy without an invasive diagnostic test. Despite the clear value of such a biomarker, whether rapid contralateral seizure spread on scalp EEG is associated with poor outcomes remains unknown.^8^

A major obstacle to answering this question is that measuring seizure spread on scalp EEG may be more challenging than on iEEG. Scalp EEG has a lower signal-to-noise ratio,^9^ and prior studies have reported modest interrater reliability for seizure and spike detection.^10,11^ The interrater agreement for seizure spread detection on scalp EEG is unknown. Furthermore, manual annotation scales poorly for research studies of large datasets,^12,13^ and no automated methods currently exist to annotate seizure spread on scalp EEG. To address this, we studied the interrater agreement of seizure onset and spread detection on scalp EEG. Also, to facilitate training and benchmarking future automated detectors of seizure onset and spread, we present the first open dataset of scalp EEG recordings with consensus annotations of both seizure onset and contralateral spread.

## MATERIALS AND METHODS

### Patient selection

We included patients with clinically diagnosed drug-resistant focal epilepsy who underwent scalp EEG recording in the Epilepsy Monitoring Unit (EMU) at the Hospital of the University of Pennsylvania (HUP) between 2017 and 2024. The eligible pool comprised 183 seizures from 54 patients with at least a seizure on scalp EEG. We targeted a cohort of 80 seizures to keep the annotation workload feasible (48 seizures per annotator). To test label consistency across seizures within the same patient, we first randomly selected 32 patients with at least two seizures and included two randomly chosen seizures from each; the remaining 16 seizures were then selected one per patient from 16 additional patients, yielding 80 seizures from 48 patients. The study protocol (#835008) was approved by the Institutional Review Board of the University of Pennsylvania with a waiver of informed consent. Clinical and demographic characteristics were obtained from the electronic medical record. SOZ localization was a consensus clinical designation made by the EMU clinical team, including at least one board-certified epileptologist, based on consistent localization across multiple habitual seizures.

### Scalp EEG recording and annotation

Scalp EEG was acquired at 256 Hz with electrodes placed according to the international 10–20 system, supplemented by inferior temporal electrodes (F9, T9, P9, F10, T10, and P10) in 45 of 48 patients; using a Natus clinical EEG system (Natus Medical, Middleton, WI, USA); an electrode posterior to Cz served as the recording reference. Recordings were archived in the native Natus format (.erd), uploaded to the IEEE portal (ieeg.org) for annotation and exported to EDF for public release. One seizure was removed as a near-duplicate of another event (separated by <1 s and electrographically indistinguishable), yielding 79 seizures from 48 patients.

Each seizure was independently annotated by three of five board-certified neurologists (EC, OF, DZ, CB, and MS; approximately 48 seizures each), similar to a multi-expert framework used for iEEG^14^. Annotators were blinded to one another’s markings and to clinical EEG reports, and attended a calibration session beforehand, in which one experienced annotator demonstrated how they identify seizure onset and contralateral spread in an example EEG. Recordings were initially displayed as referential channels; annotators switched to a preconfigured bipolar montage matching that used for clinical review, could hide selected channels, and could optionally apply a 0.5-Hz high-pass filter and a 59–61-Hz notch filter. Given the approximate event time derived from the onset marked by the treating clinicians, annotators marked (1) the seizure onset time, defined as the unequivocal electrographic onset (UEO), the first time point of a clear, continuous ictal pattern,^15^ together with all bipolar channels showing ictal activity within 1 s of the UEO; and (2) the time at which the seizure first appeared in the contralateral hemisphere, together with all involved contralateral channels at that first spread time. Annotators could indicate no seizure, no contralateral spread, or bilateral onset.

Each seizure was summarized in a six-group scheme combining onset hemisphere and contralateral spread (**Figure 1A**), from which two targets were analyzed: onset laterality (left/right/bilateral) and contralateral spread (yes/no). Bilateral onset was coded as spread = yes, with spread time equal to onset time. One clinically-annotated seizure labeled “no seizure” by two annotators was excluded, leaving 78 seizures from 48 patients (30 contributing two seizures and 18 contributing one). In 72 of these seizures, all three annotators identified a seizure; in the remaining six, one annotator labeled the event “no seizure”, so agreement was computed from the other two annotators’ labels. Midline electrodes (Fz, Cz, Pz) were excluded from laterality analyses, whereas all lateral electrodes, including inferior sub-temporal electrodes when available (N = 45 patients), were included.

**Figure 1.**
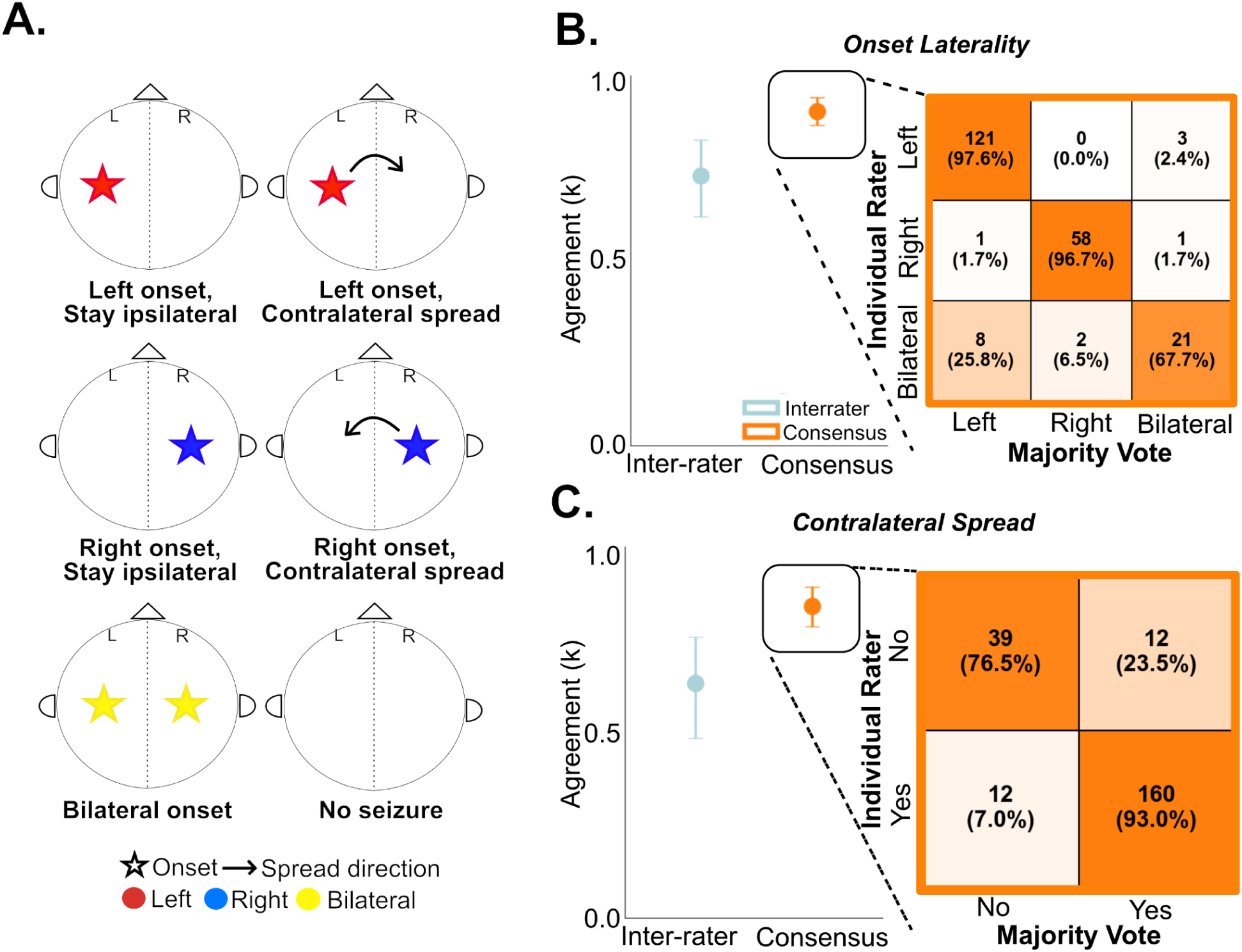
Onset and contralateral spread laterality agreement in the group. Clinicians agreed strongly on both onset laterality and the presence of contralateral spread, with near-perfect agreement against the majority-vote consensus for each. **(A)** Schematic of the six-group seizure classification scheme used to summarize spatial seizure behavior: left onset and stay ipsilateral, left onset and contralateral spread, right onset and stay ipsilateral, right onset and contralateral spread, bilateral onset, and no seizure. Stars indicate the hemisphere of onset and arrows indicate the direction of contralateral spread; the color of stars represents onset laterality (red = left, blue = right, yellow = bilateral). **(B)** Onset laterality (three categories: left, right, bilateral). Left: box-and-strip plot of Gwet k showing both inter-rater agreement (dark outline, “Interrater”) and agreement with the majority-vote consensus (orange outline, “Consensus”); black diamonds denote the mean ± linear mixed-effects model (LMM) 95% CI. Inter-rater k = 0.73 (0.62–0.83; N = 78 seizures, 48 patients); consensus k = 0.90 (0.87–0.94; N = 73 seizures). The inter-rater and consensus analyses differ in N because five seizures without a majority consensus (6.4%) were assigned no consensus label and excluded from the consensus computation, whereas all 78 seizures contribute to the inter-rater (pairwise) computation. The observed onset-category distribution used to estimate chance agreement was left 56.6% (n = 129), right 27.6% (n = 63), and bilateral 15.8% (n = 36) of individual-rater annotations. Right: consensus-only confusion matrix of individual-rater classifications (rows) against the majority-vote consensus (columns), with cell counts and row-normalized percentages; color encodes agreement (%). Individual raters classified left- and right-onset seizures highly concordantly with the majority vote (left: 121/124, 97.6%; right: 58/60, 96.7%), whereas bilateral onset was markedly less concordant (21/31, 67.7%), with individual-rater “bilateral” annotations most often corresponding to a “left” majority vote (8/31, 25.8%) rather than a “right” majority vote (2/31, 6.5%). This matrix summarizes agreement with consensus only (on the no-consensus-excluded set) and therefore does not correspond to the inter-rater component of the box-and-strip plot. **(C)** Contralateral spread existence (two categories: yes, no). Layout as in (B). Left: inter-rater k = 0.63 (0.48–0.76; N = 78 seizures) and consensus k = 0.84 (0.79–0.89; N = 75 seizures, after excluding three seizures without a majority spread consensus). The observed spread distribution was yes 76.3% (n = 174) and no 23.7% (n = 54) of individual-rater annotations. Right: consensus-only confusion matrix against the majority vote, summarizing agreement with consensus only; as in (B), the matrix reflects the consensus component, not inter-rater agreement. Individual raters agreed with the majority vote in 160/172 (93.0%) of “spread” annotations and 39/51 (76.5%) of “no spread” annotations, with the remaining discordant annotations roughly balanced between individual “no” annotations against a “spread” majority (12/172, 7.0%) and individual “spread” annotations against a “no spread” majority (12/51, 23.5%).

### Laterality agreement

Categorical agreement was quantified with the Gwet κ statistic,^16,17^ which, unlike Cohen κ, remains stable when category prevalence is highly skewed^18^ and has been used to assess interrater reliability in EEG interpretation.^19^ Gwet κ was defined as κ = (*Pa* − *Pe*)/(1 − *Pe*), where *Pa* is the observed proportion of agreement and Pe is chance agreement, estimated as [1/(*Q* − 1)] Σ π*q*(1 − π*q*) from the pooled proportion πq of ratings in each of *Q* categories. For each seizure, inter-rater Pa was the proportion of agreeing annotator pairs, and consensus *Pa* was the proportion of annotators matching the majority-vote label (a label assigned by at least two annotators); seizures without a majority label were excluded from consensus agreement only. Spread agreement was computed across all 78 seizures, without restriction to seizures with concordant onset laterality. Agreement was interpreted using conventional benchmarks:^20^ 0–0.20, slight; 0.21–0.40, fair; 0.41–0.60, moderate; 0.61–0.80, substantial; and 0.81–1.00, almost perfect. Seizure-level estimates were the mean across seizures, and patient-level estimates the mean of within-patient averages; 95% percentile bootstrap confidence intervals (CIs) were obtained from 1,000 resamples of patients.

### Channel-level agreement

Each annotator’s channel selection at onset and at spread was coded as a binary vector over up to 26 bipolar channels (24 lateral channels plus Fz–Cz and Cz–Pz); inferior sub-temporal channels were omitted when unavailable. Channel agreement was measured with Cohen κ: inter-rater agreement as the mean κ across annotator pairs, and consensus agreement as the mean κ between each annotator and the consensus channel set (channels selected by more than half of the annotators). Onset analyses included 78 seizures (76 with a consensus set where at least two experts agreed on onset laterality); spread analyses used only annotators who reported spread and required at least two such annotators per seizure (58 seizures from 37 patients; 56 with a consensus set). Annotators who reported no spread were omitted for that seizure, bilateral onset was treated as immediate spread involving all channels, and seizures without a consensus channel set were excluded from consensus agreement only. Seizure-level estimates and 95% CIs were obtained from linear mixed-effects models with a patient random intercept, and patient-level estimates from within-patient averages (mean ± 1.96 × SE). Differences between annotators were tested with ordinary least squares (OLS) regression using standard errors clustered by patient, and the annotator effect was evaluated with a Wald F-test. A linear mixed-effects model with a random intercept for patients was not used because the estimated between-patient variance was near zero and the model did not converge.

### Timing agreement

For each seizure, timing agreement was quantified as the median absolute deviation (MAD) of annotator-marked times, separately for onset and spread. Onset and spread MAD were compared within left- or right-onset seizures using the Wilcoxon signed-rank test. Seizures were also classified by the consensus structure of spread status: (1) majority (≥2/3) agreement that the seizure stayed ipsilateral; (2) majority agreement that contralateral spread occurred, for which spread MAD was computed among the agreeing annotators; and (3) complete disagreement, in which one annotator indicated no seizure and the other two disagreed on spread. Seizures in which two or more annotators indicated no seizure were excluded. Within the paired-comparison set, spread MAD was compared between seizures with unanimous (3/3) and partial (2/3) agreement on spread using the Mann–Whitney U test.^21^

### Subgroup analyses

Patients were grouped by clinical SOZ localization as temporal or non-temporal (frontal, multifocal, or unknown). At the seizure and patient levels, the between-group difference in Gwet κ (temporal − non-temporal) was estimated by resampling patients independently within each group (1,000 iterations) and considered nonsignificant when its 95% CI included zero.

### Statistical analysis and data availability

Timing measures are reported as median (interquartile range [IQR]). De-identified recordings (EDF, one file per seizure) with individual and consensus annotations are publicly available on Pennsieve;^22^ the file structure is described in the Supplementary Methods.

## RESULTS

### Patient information

Forty-eight patients were included in our analysis. **Table 1** contains a summary of the patient demographics for this study. A median of 2 seizures were obtained across patients. Thirty-two were diagnosed with temporal, six with multifocal, five with frontal, and 5 with unknown epilepsy patients. Patients underwent resection, ablation, device, or no intervention. For patients who underwent resection or ablation, 1 or 2-year Engel outcomes were determined by the board-certified neurologist treating the patient.^23^

**Table 1.** Patient demographics and clinical characteristics. The first column shows the clinical variables, the second column shows the data for the patient statistic, and the third column shows seizure statistics.

| Characteristic | Patient | Seizure |
|---|---|---|
| Total: N | 48 | 79 |
| Female: N (%) | 28 (58.3%) | 47 (59.5%) |
| Age at admission, median (range) | 38.0 (19.0-66.0) | 37.5 (19.0-66.0) |
| Seizures per patient, median (range) |  | 2.0 (1.0-2.0) |
| Localization Type |  |  |
| Temporal, N (%) | 32 (66.7%) | 56 (70.9%) |
| Multifocal: N (%) | 6 (12.5%) | 7 (8.9%) |
| Frontal: N (%) | 5 (10.4%) | 8 (10.1%) |
| Unknown, N (%) | 5 (10.4%) | 8 (10.1%) |
| Surgery performed, N (%) |  |  |
| Resection | 5 (10.4%) | 9 (11.4%) |
| Ablation | 7 (14.6%) | 10 (12.7%) |
| Device | 8 (16.7%) | 14 (17.7%) |
| No intervention | 28 (58.3%) | 46 (58.2%) |
| Engel 1 or 2 year outcome, N (%) |  |  |
| Engel class I | 7 (58.3%) | 9 (47.4%) |
| Engel class II+ | 2 (16.7%) | 4 (21.0%) |
| No outcome data available | 3 (25.0%) | 6 (31.6%) |

### Agreement on onset laterality and the presence or absence of contralateral spread was substantial

Of 79 annotated seizures, 78 were eligible for analysis (one was excluded because two annotators annotated it as “no seizure”). Across the 234 individual annotations from the 78 seizures, onset laterality was left in 55.1%, right in 26.9%, and bilateral in 15.4%; 2.1% were labeled “no seizure” and 0.4% could not be assigned a laterality (midline channels only). Onset laterality showed substantial inter-rater agreement (Gwet κ = 0.73, 95% CI = 0.62–0.83; N = 78 seizures) and almost-perfect consensus agreement (0.90, 95% CI = 0.87–0.94; N = 73 seizures, after excluding five without a majority label; **Figure 1B**), with matching patient-level estimates (0.73, 95% CI = 0.62–0.83, N = 48 patients; and 0.90, 95% CI = 0.86–0.94, N = 47; Figure S1A). Left and right annotations matched the consensus in 97.6% (121/124) and 96.7% (58/60) of cases, whereas only 67.7% (21/31) of bilateral annotations matched a bilateral consensus; the remainder corresponded more often to a left (25.8%) than a right (6.5%) majority.

Contralateral spread (yes—including bilateral onset— 76.3%, no 23.7% of annotations) showed a comparable pattern (inter-rater Gwet κ = 0.63, 95% CI = 0.48–0.76, N = 78; consensus 0.84, 95% CI = 0.79–0.89, N = 75; **Figure 1C**), with similar patient-level estimates (0.59, 95% CI = 0.43–0.72, N = 48 patients; and 0.82, 95% CI = 0.76–0.88, N = 46; Figure S1B). Among annotators who rated both seizures of the same patient (59 annotator–patient pairs from 31 patients), the same label was assigned to both seizures in 75% (44/59) of pairs for onset laterality and 78% (46/59) for contralateral spread (Figure S2). Annotators agreed with the majority in 93.0% (160/172) of “spread” and 76.5% (39/51) of “no spread” annotations. Representative cases are shown in Figures S3 and S4. Pairwise percentage agreement averaged 81.7% for onset laterality (range 66.7–93.3%) and 77.2% for contralateral spread (range 63.3–90.3%) across the 10 annotator pairs (Figure S5).

### Onset channel annotations were more reliable than spread channel annotations

At onset, channel agreement was moderate between raters (Cohen κ = 0.57, 95% CI = 0.50–0.63; N = 78 seizures/48 patients) and substantial against consensus (0.78, 95% CI = 0.75–0.82; N = 76/47; **Figure 2A**). Spread channel agreement was fair between raters (0.38, 95% CI = 0.29–0.47; N = 58/37) and substantial against consensus (0.67, 95% CI = 0.61–0.72; N = 56/37). Patient-level estimates were consistent (onset 0.56 and 0.78; spread 0.38 and 0.66; Figure S6). Annotator identity had no significant effect on consensus agreement (onset: F(4, 46) = 0.97, p = 0.432; spread: F(4, 36) = 0.26, p = 0.901; **Figure 2B**), with per-annotator means of 0.73–0.83 for onset and 0.62–0.72 for spread.

**Figure 2.**
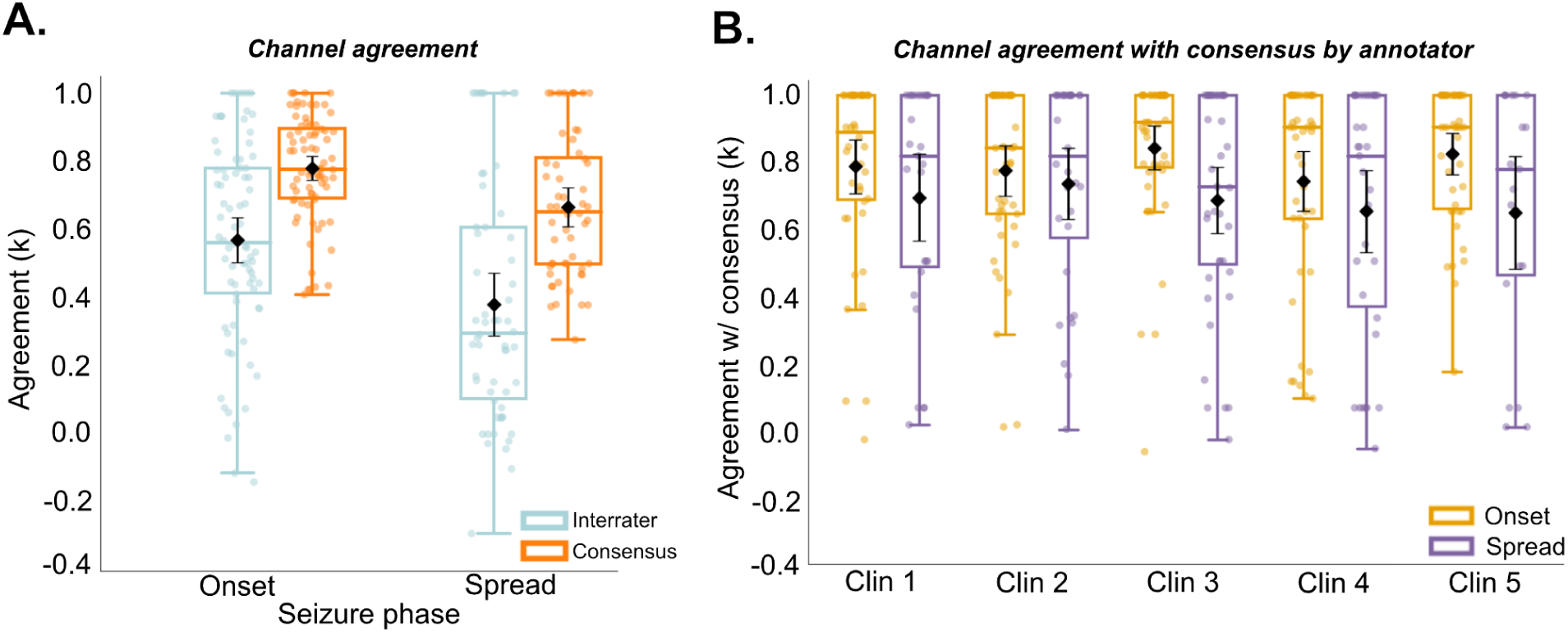
Seizure onset and contralateral spread channel annotator reliability. Channel-level annotation of which electrodes were seizing was more reliable at onset than at spread, and this reliability did not depend on which annotator was reading. **(A)** Channel-level agreement at the seizure level for onset and spread phases, measured with Cohen’s kappa (κ) from an LMM with a patient random intercept. For each phase, box-and-strip plots show inter-rater agreement (average pairwise κ across annotators; light blue outline) and agreement with the majority-vote consensus (orange outline); black diamonds denote the mean ± 95% CI. Onset channel annotations were more reliable than spread channel annotations, and for both phases agreement with consensus exceeded inter-rater agreement (onset inter-rater κ = 0.57 [0.50–0.63], N = 78 seizures / 48 patients; onset consensus κ = 0.78 [0.75–0.82], N = 76 seizures / 47 patients; spread inter-rater κ = 0.38 [0.29–0.47], N = 58 seizures / 37 patients; and spread consensus κ = 0.67 [0.61–0.72], N = 56 seizures / 37 patients). **(B)** Agreement with consensus by individual annotator (Clin 1–Clin 5) for onset (yellow) and spread (purple) channel annotations; each point is one annotator’s κ for one seizure, and black diamonds denote the mean ± 95% CI. Per-annotator mean onset κ ranged from 0.73 to 0.83 (Clin 1: 0.77, Clin 2: 0.76, Clin 3: 0.83, Clin 4: 0.73, Clin 5: 0.81), and per-annotator mean spread κ ranged from 0.62 to 0.72 (Clin 1: 0.67, Clin 2: 0.72, Clin 3: 0.66, Clin 4: 0.63, Clin 5: 0.62). Because the estimated between-patient variance in mixed-effects models was at or near zero (non-convergence), OLS models with patient-clustered robust standard errors were used and revealed no significant effect of annotator identity on agreement with consensus for either onset (F(4, 46) = 0.97, p = 0.432) or spread (F(4, 36) = 0.26, p = 0.901), indicating that annotator identity does not meaningfully alter κ values.

### Onset timing is more consistent than spread timing, but both are within seconds

Onset timing was highly consistent (median MAD 0.41 s, IQR 0.11–1.48; n = 78; **Figure 3A**). Annotators reached at least majority agreement on spread status in 75 of 78 seizures (96.2%; **Figure 3B**): in 17, the majority agreed that the seizure stayed ipsilateral (unanimous in 5, majority in 12), and in 58, that contralateral spread occurred; 54 of these had a spread MAD ≤10 s (unanimous in 44, majority in 14) and 4 had a MAD >10 s. Two of these had a MAD of 0 s because two annotators classified the seizure as bilateral onset while the third indicated no seizure. Only 3 seizures (3.8%) showed complete disagreement, with one annotator indicating no seizure and the other two disagreeing on the presence spread. In unilateral onset seizures, onset timing was more consistent than spread timing (onset median MAD 0.31 s, IQR 0.09–1.00, n = 59; spread median MAD 1.99 s, IQR 0.85–4.40, n = 42; paired Wilcoxon p < 0.001, paired n = 42; **Figure 3C**). Spread MAD did not differ significantly between seizures with unanimous and partial agreement on spread (3/3: median 1.94 s, IQR 0.73–3.67, n = 31; 2/3: median 2.95 s, IQR 1.06–6.00, n = 11; p = 0.474; Figure S7).

**Figure 3.**
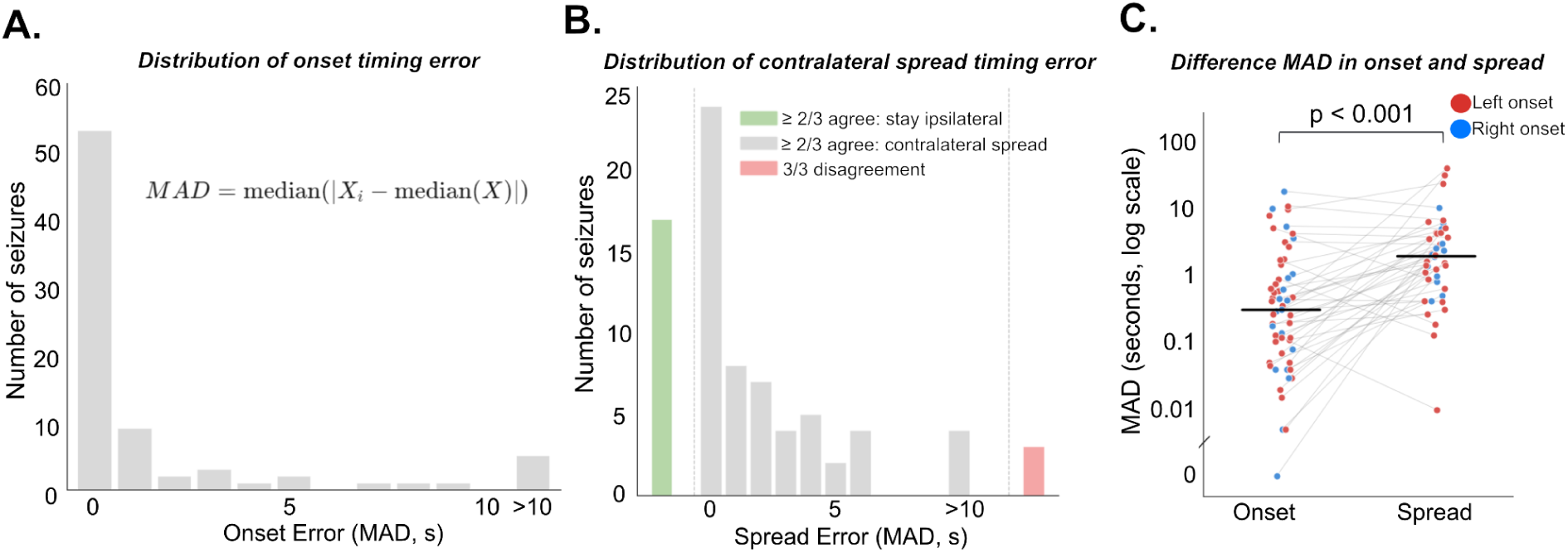
Seizure onset time and time-to-contralateral-spread annotator reliability. Onset timing was annotated with high precision, spread timing was less precise but still reasonably consistent, and most seizures reached at least majority agreement on whether spread occurred. **(A)** Distribution of onset timing error across seizures, quantified as the median absolute deviation (MAD) of annotator-marked onset times, defined as MAD = median(|Xᵢ − median(X)|) and binned in 1-s intervals with a final bin for MAD > 10 s. The distribution is strongly concentrated at low error (median onset MAD = 0.41 s, IQR 0.11–1.48; n = 78), indicating highly consistent onset timing. **(B)** Distribution of contralateral-spread timing error (MAD, 1-s bins, final bin > 10 s) across the 78 seizures, colored by agreement category: “≥2/3 agree: stay ipsilateral” (green, n = 17: unanimous 3/3 in 5 seizures, majority 2/3 in 12 seizures — plotted as a single bar to the left of 0, since no spread-timing MAD applies); “≥2/3 agree: contralateral spread” (gray, n = 58: 54 seizures with a computable spread MAD ≤ 10 s [unanimous 3/3 in 44, majority 2/3 in 14], histogrammed across the main panel, plus 4 seizures with MAD > 10 s in the rightmost bin); and “3/3 disagreement” (red, n = 3, plotted as a single bar to the right of the >10 s bin): seizures in which one annotator indicated no seizure was present while the other two disagreed with each other on whether contralateral spread occurred. Overall, 75/78 seizures (96.2%) reached at least majority (≥2/3) agreement on spread status, and complete three-way disagreement was rare (3/78, 3.8%). **(C)** Paired comparison of MAD for onset versus contralateral-spread timing among left/right-onset seizures with a computable value for both phases (each circle = one seizure), colored by consensus onset laterality (red = left onset, blue = right onset), with gray lines connecting each seizure’s onset and spread values. Bold horizontal bars indicate group medians. Onset timing showed markedly higher inter-rater reliability (lower MAD) than spread timing (onset median MAD = 0.31 s, IQR 0.09–1.00, n = 59; spread median MAD = 1.99 s, IQR 0.85–4.40, n = 42; paired Wilcoxon p < 0.001, paired n = 42). Nonetheless, spread timing remained reasonably consistent, with a median MAD of ≈ 2 s.

### Agreement was similar in temporal and non-temporal epilepsy

For onset laterality, inter-rater Gwet κ was 0.78 (95% CI = 0.64–0.89) in temporal (n = 55 seizures) and 0.62 (0.47–0.79) in non-temporal epilepsy (n = 23; difference 0.15, 95% CI = −0.05 to 0.34), and consensus κ was 0.92 (0.88–0.96; n = 52) and 0.86 (0.78–0.93; n = 21; difference 0.06, −0.03 to 0.16). For contralateral spread, inter-rater κ was 0.64 (0.47–0.80) and 0.61 (0.38–0.81; difference 0.03, −0.22 to 0.30), and consensus κ was 0.86 (0.79–0.92) and 0.80 (0.69–0.91; difference 0.05, −0.07 to 0.18; **Figure 4**). Patient-level differences showed the same pattern (onset: inter-rater 0.14, −0.08 to 0.36; consensus 0.05, −0.04 to 0.17; spread: −0.01, −0.29 to 0.28; and 0.04, −0.11 to 0.18; Figure S8). All difference CIs included zero.

**Figure 4.**
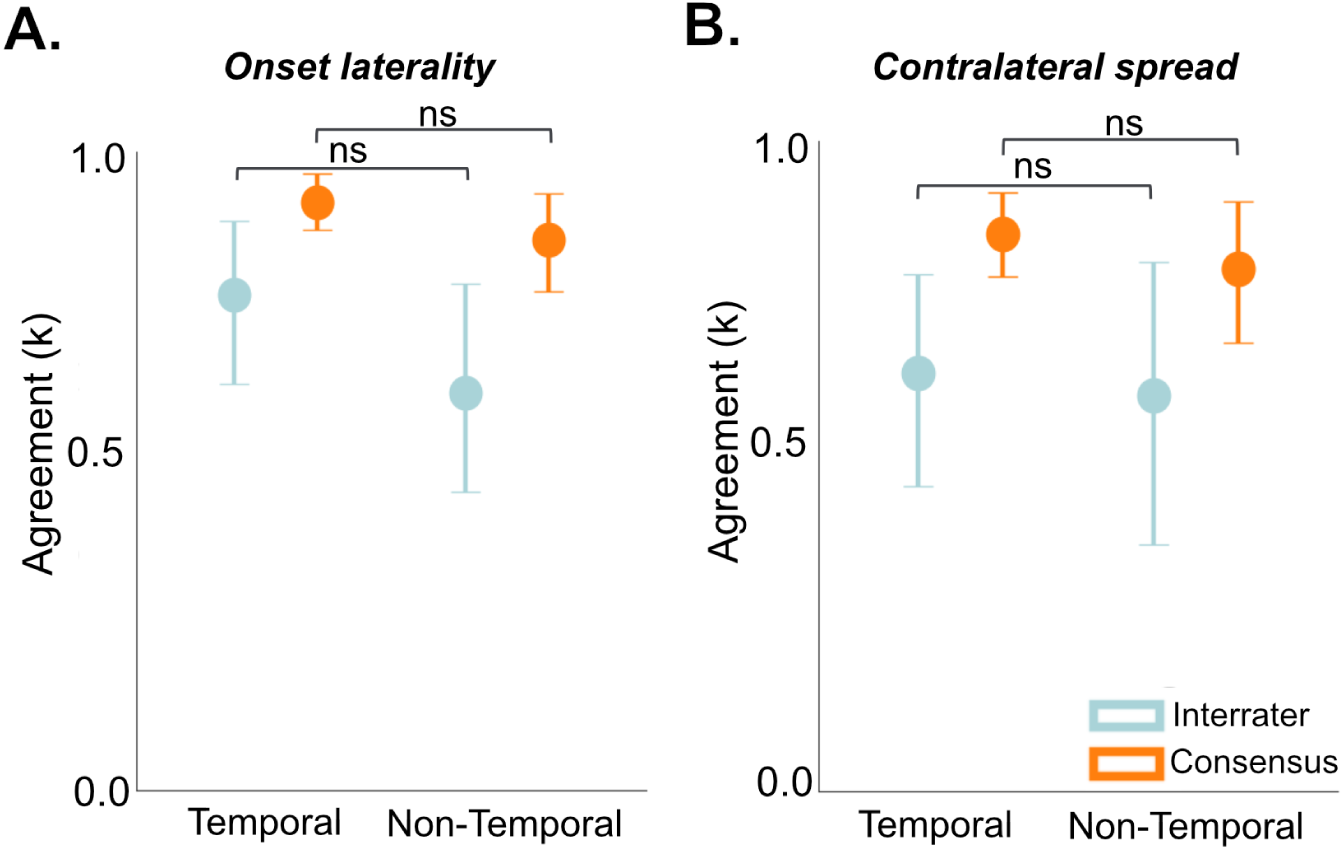
Onset-laterality and contralateral-spread agreement by epilepsy profile (temporal vs. non-temporal). Inter-rater and consensus agreement for both onset laterality and contralateral spread were similar between seizures from temporal lobe epilepsy and non-temporal lobe epilepsy. Seizure-level Gwet k for temporal lobe epilepsy versus all-other (non-temporal lobe epilepsy including multifocal, frontal, and unknown epilepsy), shown as box-and-strip plots with inter-rater agreement (light blue outline) and agreement with the majority-vote consensus (orange outline); black diamonds denote the mean ± 95% CI. **(A)** Onset laterality. Temporal lobe epilepsy: inter-rater k = 0.78 [0.66–0.89] (n = 55) and consensus k = 0.92 [0.87–0.97] (n = 52). Non-temporal lobe epilepsy: inter-rater k = 0.62 [0.40–0.84] (n = 23) and consensus k = 0.86 [0.77–0.96] (n = 21). Temporal-onset k was not significantly different from non-temporal k (Mann–Whitney U test at the seizure level, not accounting for within-patient clustering; inter-rater p = 0.111, FDR q = 0.171; consensus p = 0.128, FDR q = 0.171; “ns”). **(B)** Contralateral spread (yes/no). Temporal: inter-rater k = 0.64 [0.50–0.79] (n = 55) and consensus k = 0.86 [0.79–0.92] (n = 52). Non-temporal: inter-rater k = 0.61 [0.40–0.81] (n = 23) and consensus k = 0.80 [0.70–0.91] (n = 23). k for temporal lobe epilepsy was not significantly different from non-temporal lobe epilepsy (inter-rater p = 0.753, FDR q = 0.990; consensus p = 0.893, FDR q = 0.990; “ns”). FDR q-values were computed across the four comparisons shown here.

## DISCUSSION

Using a standardized protocol in which each seizure was reviewed by three of five epileptologists, we found substantial agreement on the laterality of seizure onset and on the presence and timing of contralateral spread on scalp EEG. Agreement was consistently higher for onset than for spread and higher for consensus than inter-rater agreement, and it did not differ significantly across annotators or between temporal and non-temporal epilepsy. These results suggest that seizure spread on scalp EEG can be annotated reliably, supporting the feasibility of measuring spread speed on scalp EEG as a potential biomarker of diffuse epileptic networks.

### There was strong agreement about onset laterality

When annotators labeled a seizure as unilateral, they rarely disagreed on the side: left and right annotations matched the consensus in more than 96% of cases. Almost all disagreement involved bilateral onset, which matched a bilateral consensus in only 67.7% of annotations and was otherwise more often assigned a left than a right majority. This reflects the difficulty of deciding from scalp EEG alone whether bihemispheric ictal activity represents genuinely bilateral onset or rapid propagation from a unilateral focus. This distinction often remains unresolved even after invasive evaluation: among patients with bilateral independent or unclear seizure onset on scalp EEG, stereo-EEG identified a single SOZ in only 14%.^24^

### Clinicians agreed on the presence of contralateral spread, but not the specific channels involved

Agreement on whether contralateral spread occurred was also strong (inter-rater Gwet κ = 0.63; consensus 0.84), though slightly lower than onset laterality. On scalp EEG, contralateral ictal activity may represent genuine spread, volume conduction, or an independent bilateral onset. Reviews correlating invasive and scalp recordings attribute this ambiguity to the limited spatial resolution and volume-conduction properties intrinsic to extracranial recording.^25^ Spread channels are also typically lower in amplitude, more spatially distributed, and more affected by overlapping propagated and volume-conducted fields than the focal activity at onset. Even so, annotators interpreted this ambiguity consistently in most seizures, although agreement does not establish that their interpretation reflects true propagation. Our inter-rater onset and spread channel agreements were lower than those reported in a study of iEEG (0.57 vs 0.64 for onset 0.38 vs. 0.62 for spread channels).^14^ The lower interrater reliability for onset and spread channels on scalp EEG potentially reflect the higher signal-to-noise ratio in iEEG. Also, the drop in interrater reliability from onset to spread channels was higher in scalp EEG, which may reflect the greater confusion about spread vs. volume conduction in scalp EEG. However, the interpretation of this cross-modality comparison is limited as a non-simultaneous study of the same seizures or patients with scalp EEG and iEEG remains an open area of investigation.

### Onset and spread timing were consistent

Onset timing varied by a median of 0.41 s across annotators, and spread timing, although less consistent, by 1.99 s. This is within the windows typically used to define early spread for prognostic purposes: iEEG studies correlating spread timing with surgical outcome have used early-spread cutoffs from 1 to 14 s, with a systematic review finding that thresholds of 3 to 10 seconds have the strongest associations with postoperative seizure freedom.^3,5^ When clinicians agree that contralateral spread occurred, the variability in their spread-time estimates is within these cutoffs (although could lead to discordance for the more conservative 3 s cutoff). Spread-timing variability was also similar whether two or all three annotators agreed that spread occurred, suggesting that even when experts disagree on *whether* spread has occurred, those who identified spread agreed on *when* it occurred.

### Toward automated spread detection

Whether the speed of seizure spread on scalp EEG predicts surgical outcome has not been established and requires large multicenter cohorts of surgical patients, to which manual annotation cannot scale. Our public dataset, which pairs each seizure with three independent expert annotations of onset and spread times and channels together with a majority-vote consensus, supports the development and validation of automated algorithms that could make such studies feasible. Such algorithms could also provide a reproducible measure of spread that does not depend on individual readers. Also, that algorithm could give better spatiotemporal resolution than humans.

### Limitations

This study has several limitations. First, it was conducted at a single center and may underestimate the variability in annotation across centers with different training and reading conventions. Second, most patients had temporal lobe epilepsy; agreement did not differ significantly between temporal and non-temporal epilepsy, but the non-temporal subgroup was small, so the generalizability of these estimates to extratemporal epilepsy remains uncertain. Third, the majority-vote consensus of three annotators is a reference standard, and we did not validate scalp EEG spread against intracranial recordings. Fourth, each seizure was reviewed by three of five annotators rather than by all five, so annotator pairs shared different subsets of seizures. Finally, we focused our analysis on contralateral seizure spread, rather than spread within a hemisphere, given evidence that scalp EEG is primarily efficacious for lateralizing epilepsy.^26^ Our finding of lower interrater reliability for annotating the specific channels involved at contralateral seizure spread support that measuring intrahemispheric seizure spread may be less feasible on scalp EEG, but future studies will need to directly test this hypothesis.

### Conclusion

Epileptologists showed substantial agreement on seizure onset laterality and on whether and when seizures spread to the contralateral hemisphere on scalp EEG. These findings support the plausibility of using scalp EEG spread to assess focal versus diffuse seizure networks, as has been done with intracranial EEG, and warrant larger multicenter studies measuring the association in spread speed with surgical outcome.

## DATA AVAILABILITY STATEMENT

All code used to perform analyses is publicly available at https://github.com/penn-cnt/scalp_seizure_annotation. The data can be downloaded from pennsieve.io and can be referenced here: https://doi.org/10.26275/A3ND-UOC9. EDF data can be streamed on Pennsieve. Details on data download and API to interact with the data are previously described^22^ and on the Pennseive documentation hub (https://docs.pennsieve.io/docs/getting-started).

