## Supplementary materials for "Interrater agreement of seizure onset and spread in scalp EEG"

**SUPPLEMENTARY MATERIALS**
***Supplemental methods***

**Data Organization**
At the EEG data level, file names use unique identifiers (PennEPI[N]_seizure[M].edf). Files also include patient-level demographic data (Participants_PennEPIID.csv) and seizure-level annotations (Annotation_PennEPIID.csv).

**Participants_PennEPIID.csv**
This table contains participant-level metadata and is presented in Table 1. Each row represents a unique participant identified by "PennEPI". To match the edf file and annotation, seizure_number is identified. Demographic variables include biological "gender" (F: female, M: male) and "age" at admission (in years). Clinical characterization variables describe epilepsy subtypes with “scalp_localization” (temporal, multifocal, frontal, and unknown) and surgery performed with “Surgery received” (resection, laser ablation, VNS, RNS, DBS, and no intervention received). Surgical outcome is described by the "Engel" field, which contains the Engel score for post-surgical seizure freedom assessed at 12 or 24 months post-surgery.

**Annotation_PennEPIID.csv**
This table contains seizure-level metadata describing both seizure characteristics and expert clinician annotations. Each row represents a single seizure event with annotations from three expert clinicians. Seizure identification fields include "PennEPI" and "seizure_number". For the edf file name, “edf_file_name” is listed. For each seizure, clinicians independently marked the unequivocal electrographic onset (UEO) time and the channels exhibiting seizure activity at onset and at spread time to contralateral hemisphere. Each annotations are stored in list format: "Your_onset_time (s)_[N]" contains each clinician's marked onset time (in seconds) and "Your_onset_channel_[N]" contains the bipolar channel pairs that each clinician identifies as seizing at onset. If the seizure appears to begin diffusely, they mark diffuse or all channel pairs (N=1 to 5). "Your_contralateral_spread_time (s)_[N]" contains each clinicians mark the time (in seconds) that the seizure appears to first spread to the opposite hemisphere. If this never happens, or if the clinician marks bilateral onset channels, it is NaN. "Your_contralateral_spread_channel_[N]" contains the channels each clinician identifies seizure spread to the opposite side. To find out where seizure off, it is listed as “seizure_off” and to the point when edf file recording off, it is listed as “recording_off”. The “Group_[N]” field contains the group based on each annotator’s marked onset and spread. We included left and right hemispheric channel pairs (midline was excluded) when we calculated it in Group_[N] (left onset and ipsilateral spread, left onset and contralateral spread, right onset and ipsilateral spread, right onset and contralateral

spread, bilateral onset, no seizure, and other); other is the category when onset has only midline, for example, onset starts at the midline and spreads to the left side. The "Majority_Group" field contains the consensus group determined by majority voting across individual clinician annotations. The “Agreed_Annotators” field contains which annotator [N] was involved in the Majority_Group.

**scalp EEG**

Each EEG file is named by the patient identifier (PennEPI) and seizure number (e.g., PennEPI100_seizure29.edf). Both identifiers can be matched with Annotations_PennEPIID.csv and Participants_PennEPIID.csv. Each EDF file contains the EEG from 60 seconds before the earliest annotated seizure onset to 60 seconds after the latest annotated seizure offset. When fewer than 60 seconds were available before the onset or after the offset, the maximum available duration was included. Because recording lengths differ across EDF files, Annotations_PennEPIID.csv also reports seizure_off and recording_off. Each EDF file contains the sampling frequency, channel names, and recording duration. To avoid introducing artifacts or degrading information in the raw signal, no signal preprocessing was applied to the EEG data.

*Supplemental figures*

**Fig. S1. Patient-level agreement for onset laterality and contralateral spread.**

Agreement was estimated with Gwet's k (shown as κ) at the patient level. Of 79 annotated seizures from 48 patients, one seizure was excluded because two or more annotators labeled it "no seizure," leaving 78 seizures (48 patients; 30 with two seizures and 18 with one). Each seizure was reviewed by 3 of the 5 annotators, and 72 seizures had three valid ratings and six had two. For each seizure, observed agreement (p_o) was the proportion of annotator pairs giving the same label for inter-rater agreement (light blue), and the proportion of annotators whose label matched the majority label (at least two annotators agreeing) for consensus agreement (orange). Seizures without a majority label were excluded from the consensus estimates. Points show the mean across patients, and error bars show 95% confidence intervals from 1,000 bootstrap resamples of patients. **(A)** Onset laterality (Left, Right, Bilateral). Labels were distributed as Left 56.6%, Right 27.6%, and Bilateral 15.8%. Inter-rater agreement was κ = 0.73 (95% CI 0.62–0.83; 48 patients), and consensus agreement was κ = 0.90 (95% CI 0.86–0.94; 47 patients, 73 seizures, after excluding 5 seizures without a majority label). **(B)** Contralateral spread (Yes/No), with bilateral onset counted as Yes. Labels were distributed as Yes 76.3% and No 23.7%. Inter-rater agreement was κ = 0.59 (95% CI 0.43–0.72; 48 patients), and consensus agreement was κ = 0.82 (95% CI 0.76–0.88; 46 patients, 75 seizures, after excluding 3 seizures without a majority label).

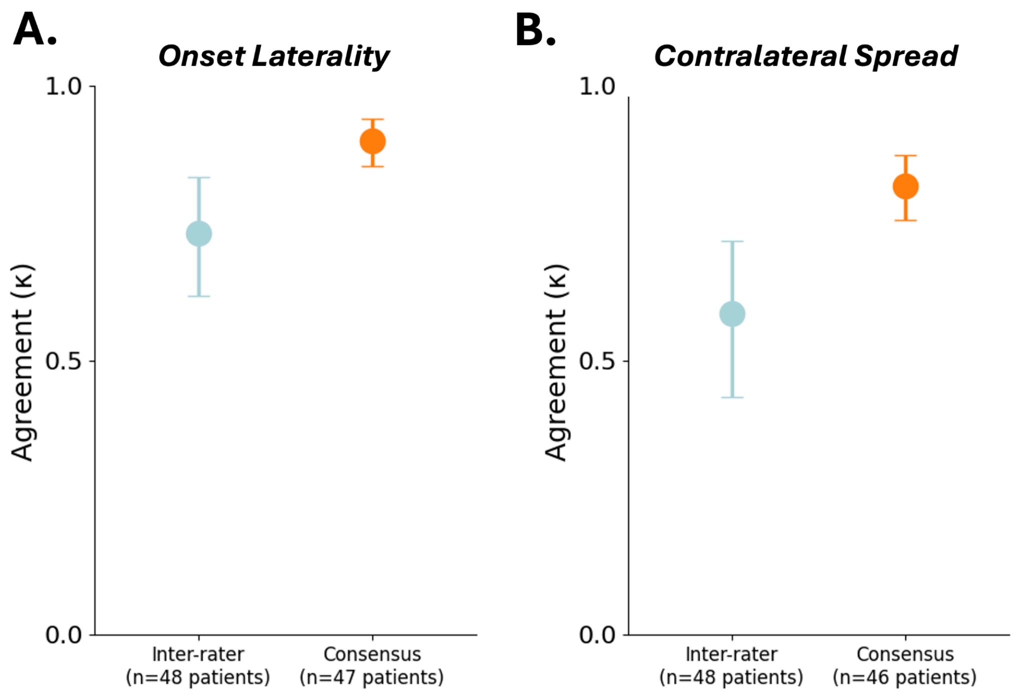

**Figure S2. Consistency of each annotator's labels across the two seizures of the same patient.**

Labels for (A) onset laterality and (B) contralateral spread given by the same annotator to the two seizures of patients who contributed two seizures (31 patients). Because annotators were assigned independently for each seizure, only annotators who rated both seizures of a patient are included. One annotator rated both seizures in 9 patients, two annotators in 16, and all three in 6, giving 59 annotator–patient pairs. Each circle is one annotator–patient pair. The x-axis shows the label for seizure 1 and the y-axis the label for seizure 2. Dashed boxes mark pairs in which the annotator gave the same label to both seizures. "No seizure" indicates that the annotator judged the event not to be a seizure. The same label was given to both seizures in 44 of 59 pairs (75%) for onset laterality and in 46 of 59 pairs (78%) for contralateral spread.

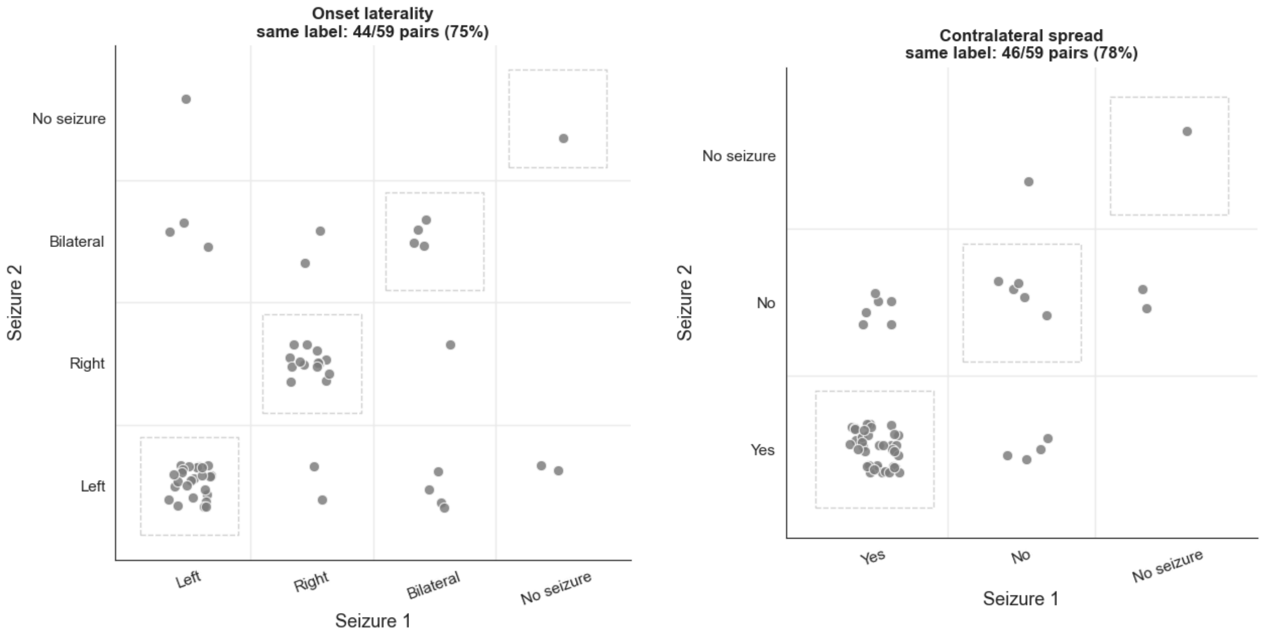

**Fig. S3. Representative case with unanimous onset laterality agreement (3/3) and majority agreement on contralateral spread (2/3).**

Representative scalp EEG recording with time on the x-axis and channel pairs on the y-axis, shown as reviewer's arrow. Left onset with contralateral spread, demonstrating ictal activity beginning over left-hemispheric channels. Two out of three experts annotated that the seizure subsequently spread to the right-hemispheric channels, and one reviewer annotated that there was no contralateral spread.

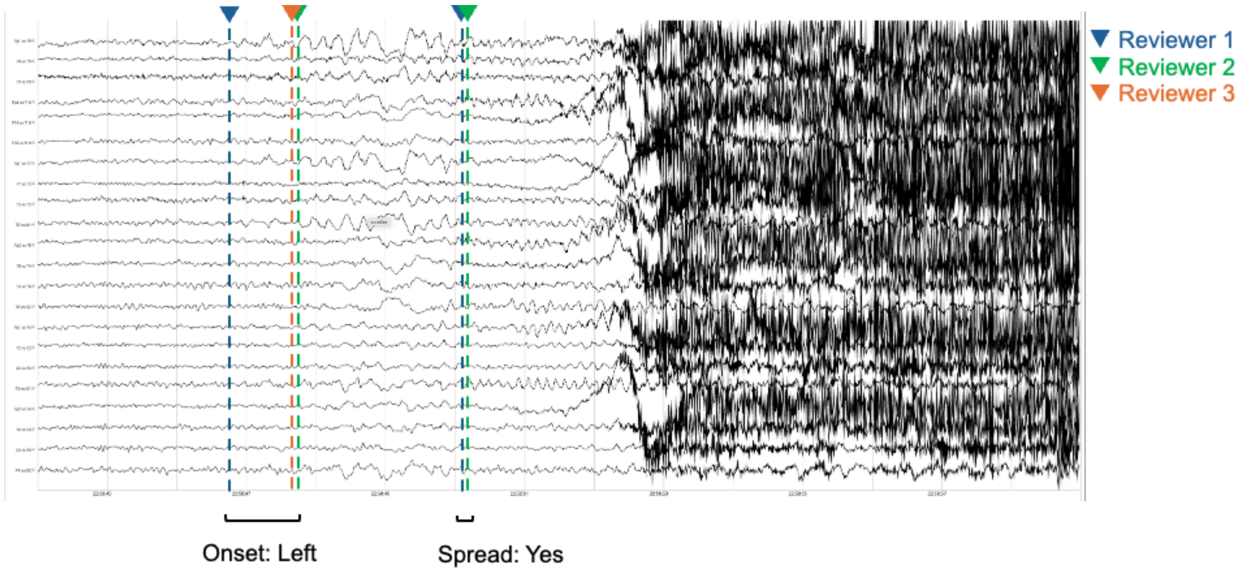

**Fig. S4. Representative scalp EEG example of inter-rater disagreement in seizure-grouping classification, illustrating the electrographic ambiguity underlying discordant annotations.**
Representative scalp EEG recording with time on the x-axis and channel pairs on the y-axis, shown as individual line traces, illustrating a case in which three clinician annotators assigned discordant seizure-grouping labels based on onset and spread. The three annotators differed in grouping categories based on onset and spread. Reviewer 1 classified the seizure as *left onset, stay ipsilateral* (without spread); Reviewer 2 as *bilateral onset*; and Reviewer 3 as *right onset, contralateral spread*. This case illustrates compounded disagreement, in which uncertainty about when the seizure begins is accompanied by uncertainty about where it begins and whether it spreads.

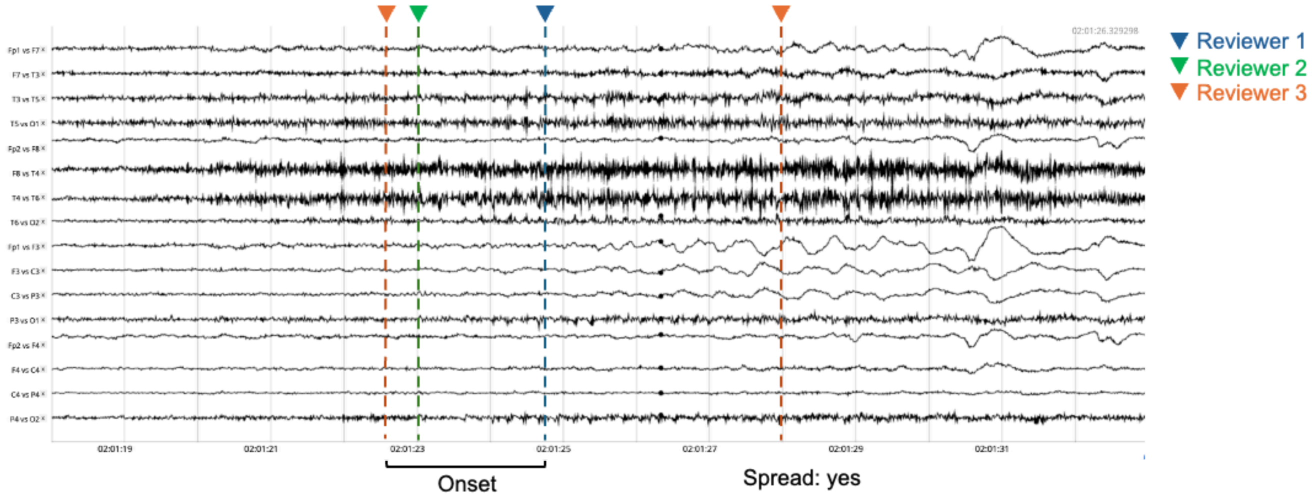

**Fig. S5. Pairwise inter-rater agreement for onset laterality and contralateral spread.**

**(A) Onset laterality.** *Left*, contingency tables for all 10 annotator pairs (C(5,2)). Each seizure was reviewed by 3 of the 5 annotators, so each table includes only the seizures that both annotators in that pair reviewed (n = 13–31 seizures per pair). Rows give the first annotator's label and columns the second annotator's label (Left, Right, Bilateral). The Total row and column (gray) show how often each annotator used each category. *Right*, 5 × 5 percentage agreement matrix. Each cell shows the percentage of shared seizures that both annotators assigned the same label, with the number of shared seizures in parentheses. Darker shading means lower agreement (scale 0–100%), and the diagonal is left blank. Mean pairwise agreement was 81.7% (range 66.7–93.3%).

**(B) Contralateral spread.** Same layout as (A), using a binary label (Yes/No). Seizures labeled as bilateral onset were counted as spread = Yes. Mean pairwise agreement was 77.2% (range 63.3–90.3%). Percentage agreement is raw observed agreement and is not corrected for chance. Across all 229 ratings (79 seizures, 48 patients), labels were unevenly distributed: Left 56.3%, Right 27.5%, Bilateral 16.2%, and spread Yes 76.4% vs No 23.6%.

**A.**

**Onset laterality — pairwise annotator agreement table (3×3, 10 pairs, with totals)**

Annotator 1 vs 2 (n=29)

| | Left | Right | Bilateral | Total |
|---|---|---|---|---|
| Left | 16 | 2 | 0 | 18 |
| Right | 0 | 7 | 0 | 7 |
| Bilateral | 1 | 2 | 1 | 4 |
| Total | 17 | 11 | 1 | 29 |

Annotator 1 vs 3 (n=15)

| | Left | Right | Bilateral | Total |
|---|---|---|---|---|
| Left | 7 | 0 | 2 | 9 |
| Right | 0 | 4 | 0 | 4 |
| Bilateral | 1 | 0 | 1 | 2 |
| Total | 8 | 4 | 3 | 15 |

Annotator 1 vs 4 (n=15)

| | Left | Right | Bilateral | Total |
|---|---|---|---|---|
| Left | 7 | 1 | 3 | 11 |
| Right | 0 | 3 | 0 | 3 |
| Bilateral | 1 | 0 | 0 | 1 |
| Total | 8 | 4 | 3 | 15 |

Annotator 1 vs 5 (n=29)

| | Left | Right | Bilateral | Total |
|---|---|---|---|---|
| Left | 19 | 0 | 1 | 20 |
| Right | 0 | 6 | 0 | 6 |
| Bilateral | 2 | 1 | 0 | 3 |
| Total | 21 | 7 | 1 | 29 |

Annotator 2 vs 3 (n=31)

| | Left | Right | Bilateral | Total |
|---|---|---|---|---|
| Left | 15 | 0 | 1 | 16 |
| Right | 1 | 8 | 1 | 10 |
| Bilateral | 1 | 0 | 4 | 5 |
| Total | 17 | 8 | 6 | 31 |

Annotator 2 vs 4 (n=15)

| | Left | Right | Bilateral | Total |
|---|---|---|---|---|
| Left | 6 | 0 | 2 | 8 |
| Right | 0 | 4 | 0 | 4 |
| Bilateral | 0 | 0 | 3 | 3 |
| Total | 6 | 4 | 5 | 15 |

Annotator 2 vs 5 (n=15)

| | Left | Right | Bilateral | Total |
|---|---|---|---|---|
| Left | 10 | 0 | 0 | 10 |
| Right | 1 | 4 | 0 | 5 |
| Bilateral | 0 | 0 | 0 | 0 |
| Total | 11 | 4 | 0 | 15 |

Annotator 3 vs 4 (n=30)

| | Left | Right | Bilateral | Total |
|---|---|---|---|---|
| Left | 12 | 0 | 3 | 15 |
| Right | 0 | 8 | 1 | 9 |
| Bilateral | 1 | 0 | 5 | 6 |
| Total | 13 | 8 | 9 | 30 |

Annotator 3 vs 5 (n=13)

| | Left | Right | Bilateral | Total |
|---|---|---|---|---|
| Left | 5 | 0 | 0 | 5 |
| Right | 0 | 5 | 0 | 5 |
| Bilateral | 1 | 1 | 1 | 3 |
| Total | 6 | 6 | 1 | 13 |

Annotator 4 vs 5 (n=30)

| | Left | Right | Bilateral | Total |
|---|---|---|---|---|
| Left | 13 | 0 | 1 | 14 |
| Right | 0 | 7 | 1 | 8 |
| Bilateral | 6 | 2 | 0 | 8 |
| Total | 19 | 9 | 2 | 30 |

**Onset laterality — Percentage agreement (5×5)**

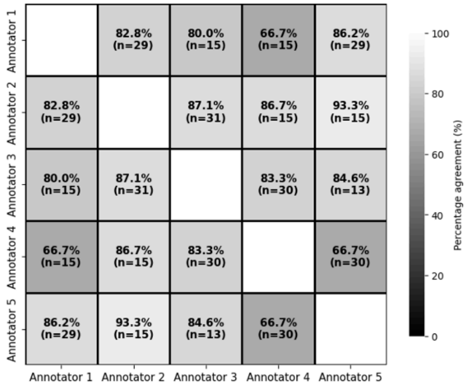


**B.**

**Contralateral spread (Yes/No) — pairwise annotator agreement table (2×2, 10 pairs, with totals)**

Annotator 1 vs 2 (n=29)

| | Yes | No | Total |
|---|---|---|---|
| Yes | 19 | 1 | 20 |
| No | 5 | 4 | 9 |
| Total | 24 | 5 | 29 |

Annotator 1 vs 3 (n=15)

| | Yes | No | Total |
|---|---|---|---|
| Yes | 13 | 1 | 14 |
| No | 1 | 0 | 1 |
| Total | 14 | 1 | 15 |

Annotator 1 vs 4 (n=15)

| | Yes | No | Total |
|---|---|---|---|
| Yes | 9 | 0 | 9 |
| No | 5 | 1 | 6 |
| Total | 14 | 1 | 15 |

Annotator 1 vs 5 (n=29)

| | Yes | No | Total |
|---|---|---|---|
| Yes | 11 | 4 | 15 |
| No | 3 | 11 | 14 |
| Total | 14 | 15 | 29 |

Annotator 2 vs 3 (n=31)

| | Yes | No | Total |
|---|---|---|---|
| Yes | 25 | 2 | 27 |
| No | 1 | 3 | 4 |
| Total | 26 | 5 | 31 |

Annotator 2 vs 4 (n=15)

| | Yes | No | Total |
|---|---|---|---|
| Yes | 11 | 0 | 11 |
| No | 3 | 1 | 4 |
| Total | 14 | 1 | 15 |

Annotator 2 vs 5 (n=15)

| | Yes | No | Total |
|---|---|---|---|
| Yes | 5 | 4 | 9 |
| No | 1 | 5 | 6 |
| Total | 6 | 9 | 15 |

Annotator 3 vs 4 (n=30)

| | Yes | No | Total |
|---|---|---|---|
| Yes | 25 | 0 | 25 |
| No | 4 | 1 | 5 |
| Total | 29 | 1 | 30 |

Annotator 3 vs 5 (n=13)

| | Yes | No | Total |
|---|---|---|---|
| Yes | 10 | 3 | 13 |
| No | 0 | 0 | 0 |
| Total | 10 | 3 | 13 |

Annotator 4 vs 5 (n=30)

| | Yes | No | Total |
|---|---|---|---|
| Yes | 18 | 11 | 29 |
| No | 0 | 1 | 1 |
| Total | 18 | 12 | 30 |

**Contralateral spread (Yes/No) — Percentage agreement (5×5)**

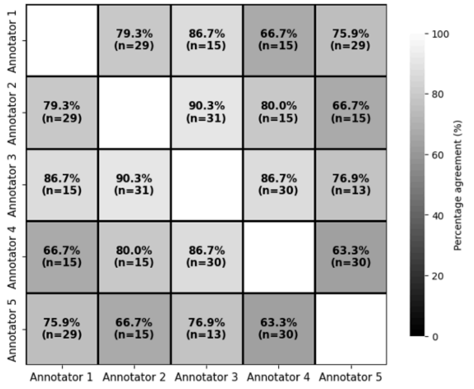

**Fig. S6. Patient-level channel agreement for seizure onset and contralateral spread.**
This figure shows the patient-level counterpart of Figure 2A. Agreement on the EEG channels marked at seizure onset and at contralateral spread was quantified with Cohen's κ. For each seizure, inter-rater agreement (light blue) was the mean κ across all pairs of annotators, and consensus agreement (orange) was the mean κ between each annotator and the consensus channel set. The consensus set contained the channels selected by more than half of the annotators. Seizures in which no channel met this criterion had no consensus set and were excluded from the consensus estimate only. Onset agreement used the annotators with a valid onset annotation in each seizure (78 seizures from 48 patients). Spread agreement used only the annotators who reported contralateral spread, and required at least two such annotators per seizure (58 seizures from 37 patients). Annotators who reported no spread were left out of the comparison for that seizure, and a bilateral onset was treated as immediate spread to all channels. Seizure-level κ values were averaged within each patient, so each point represents one patient. Boxes show the median and interquartile range, and whiskers extend to 1.5 times the interquartile range. Black diamonds show the mean across patients with 95% confidence intervals (mean ± 1.96 × SE). Onset inter-rater agreement was κ = 0.56 (95% CI 0.50–0.63; 48 patients), and onset consensus agreement was κ = 0.78 (0.74–0.81; 47 patients). Spread inter-rater agreement was κ = 0.38 (0.29–0.47; 37 patients), and spread consensus agreement was κ = 0.66 (0.60–0.73; 37 patients). These patient-level estimates closely match the seizure-level estimates in Figure 2A (onset 0.57 and 0.78; spread 0.38 and 0.67).

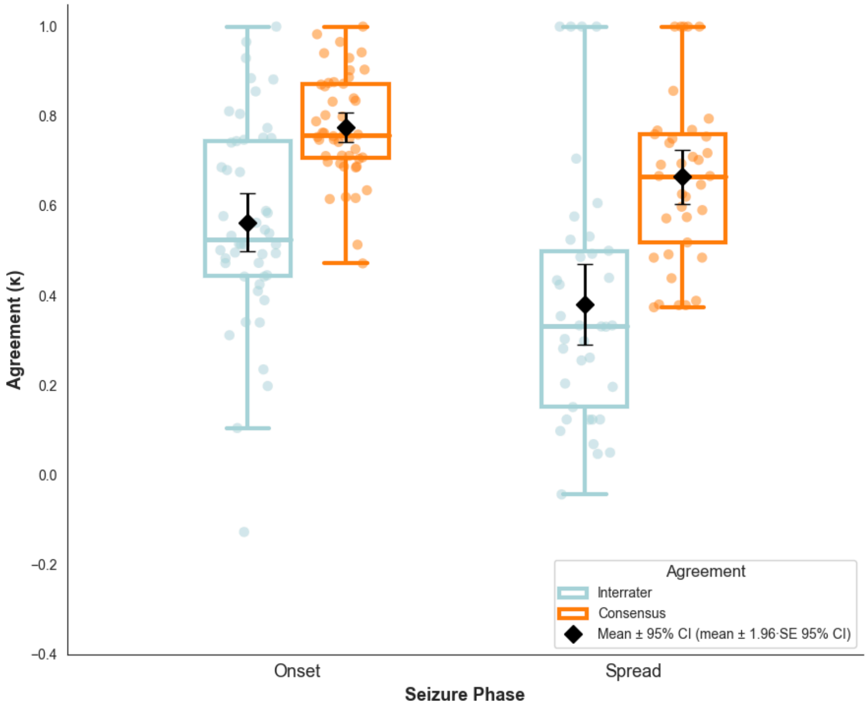

**Fig. S7. Relationship between annotator agreement and spread-timing error.** Seizures with only partial agreement on spread existence (2/3) showed higher spread-timing MAD among the two agreeing annotators than those with unanimous agreement (3/3: median MAD = 1.94 s, IQR 0.73–3.67, n = 31; 2/3: median MAD = 2.95 s, IQR 1.06–6.00, n = 11). This difference did not reach statistical significance (Mann–Whitney p = 0.474). This analysis also documents that a spread-timing MAD can be computed whenever at least two of three annotators agree spread occurred, as used in Figure 3C.

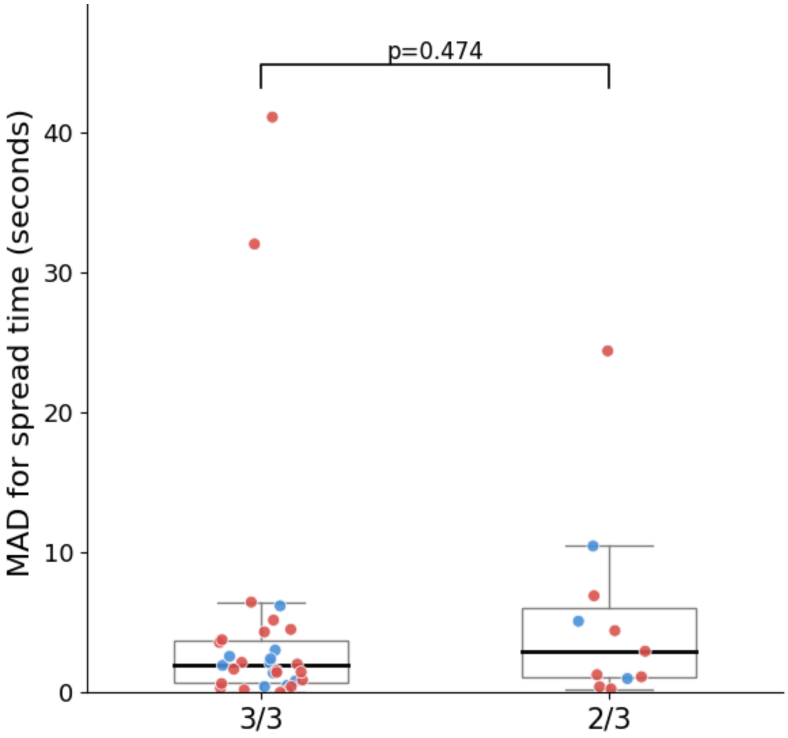

**Fig S8. Patient-level onset-laterality and contralateral-spread agreement by seizure localization (temporal vs. non-temporal).**

Agreement was estimated with Gwet's k (shown as κ) separately for seizures with temporal scalp localization (Temporal; 32 patients) and all other seizures (Non-Temporal; 16 patients, comprising frontal, multifocal, and unknown localization). Localization was assigned per seizure, and no patient had seizures in both groups. Of 79 annotated seizures, one seizure labeled "no seizure" by two or more annotators was excluded, leaving 55 temporal and 23 non-temporal seizures. Each seizure was reviewed by 3 of the 5 annotators. Seizure-level k values were averaged within each patient, so that each patient contributed equally. Points show the mean across patients, and error bars show 95% confidence intervals from 1,000 bootstrap resamples of patients. The difference between groups (Temporal − Non-Temporal) was estimated by resampling patients within each group, and was considered non-significant when its 95% confidence interval included zero. **(A)** Onset laterality (Left, Right, Bilateral). Inter-rater agreement was κ = 0.78 (95% CI 0.64–0.89; 32 patients) for temporal and κ = 0.64 (0.46–0.82; 16 patients) for non-temporal seizures, a difference of 0.14 (95% CI −0.08 to 0.36). Consensus agreement was κ = 0.92 (0.87–0.96; 31 patients, 52 seizures) and κ = 0.86 (0.77–0.94; 16 patients, 21 seizures), respectively, a difference of 0.05 (−0.04 to 0.17). **(B)** Contralateral spread (Yes/No), with bilateral onset counted as Yes. Inter-rater agreement was κ = 0.59 (0.40–0.75; 32 patients) for temporal and κ = 0.59 (0.37–0.81; 16 patients) for non-temporal seizures, a difference of −0.01 (−0.29 to 0.28). Consensus agreement was κ = 0.83 (0.76–0.91; 30 patients, 52 seizures) and κ = 0.80 (0.69–0.91; 16 patients, 23 seizures), respectively, a difference of 0.04 (−0.11 to 0.18).

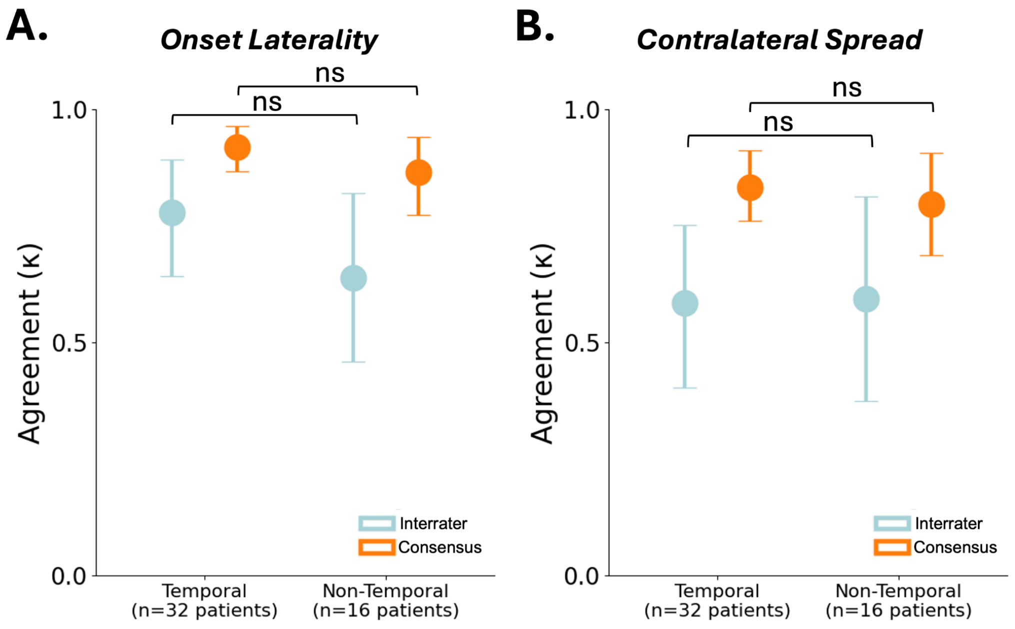
